# VACS 2.0 Frailty Index Correlates with Soluble TNF Receptor Levels in Aging Veterans

**DOI:** 10.64898/2026.05.24.26353987

**Authors:** Sarah Carbone, Brigid Wilson, Corinne Kowal, Teresa Dolinar, Kostadinova Lenche, Donald D. Anthony, Carey L. Shive

## Abstract

The VACS 2.0 Frailty Index was developed using the VA health records system to identify frailty and predict mortality in older Veterans that were living with HIV. Systemic inflammatory indices have been associated with frailty, but little is known about the association between frailty and immunosenescence. We aim to investigate the potential link between soluble inflammatory indices, T cell expression of exhaustion and senescence markers, and frailty as measured by the VACS 2.0 index. We analyzed a one-time blood draw for plasma levels of inflammatory indices, T cell subsets and expression of exhaustion and senescence markers, and calculated VACS 2.0 index scores in a cohort of 30 older (≥65 years) Veteran participants. We found that VACS 2.0 scores correlated with the number of prescribed medications in the older Veterans. Soluble TNF receptor levels strongly correlated with VACS 2.0 frailty scores. How these soluble TNF receptors are generated and whether they mechanistically contribute to frailty warrants further investigation.

## BACKGROUND

Frailty syndrome is a clinical state of increased vulnerability and decreased resilience to stressors that is associated with aging and a decline in physiological function^1^. Frailty often results in increased hospitalization, falls, and mortality^2^. A 2015 study examining frailty in the non-nursing home US population aged 65 and older, found that 15% were frail and 45% were pre-frail based on measurements using the Fried frailty assessment^3^. A more recent systematic review of global frailty (62 countries) found 12% frailty and 46% pre-frailty in studies using physical frailty measures like the Fried frailty assessment, and 24% frailty and 49% pre-frailty in studies that used frailty indexes^4^. Although life expectancy has steadily increased over the last 50 years, health-span (years of good health) has not kept pace. In 2024, approximately 18% of the US population was over 65 years old, by the year 2050, it is predicted that this same age group will increase to nearly 22% of the US population, making frailty an even greater health issue for our society. The Veteran Aging Cohort Study (VACS) 2.0 frailty index was developed using a Veteran HIV patient cohort. It estimates risk of 5-year all-cause mortality in patients with HIV and the greater the score, the higher the probability of frailty^5^. More recent studies have since validated the VACS 2.0 frailty index in aging populations without HIV^6, 7^.

In general, frailty indexes rely on health records to define frailty based on declining function, weight loss, diagnosed morbidities, cognition and mood, chronic pain, and sensory loss, with no patient interaction at the time of assessment. A frailty score is then calculated based on clinical data gathered from the health records^8^. Clinical health data used to generate the VACS 2.0 frailty score include: chronological age, sex, body mass index (BMI), CD4+ cell count, HIV-1 RNA viral load, hepatitis C virus (HCV) co-infection status, estimated glomerular filtration rate (eGFR), hemoglobin, alanine transaminase (ALT), aspartate aminotransferase (AST), platelet count, white blood cell (WBC) count, and albumin level (g/dL)^9^.

A frailty phenotype, on the other hand, is generated based on an active physical assessment or measurement of abilities. The best-known validated frailty phenotype assessment is the Fried Frailty Score^1^. The Fried Frailty score is based on five criteria; 1) unintentional weight loss of 10 or more pounds within the past year, 2) low physical activity, 3) exhaustion (both measured with a self-reported questionnaire), 4) slow walking gait measured by a timed distance, and 5) weakness as measured by grip strength^1^. Unlike many frailty indexes, the Fried Frailty Score can be used to categorize patients as frail (≥3 criteria), pre-frail (1-2 criteria), or robust/not frail (0 criteria)^1^. Aging is often accompanied by increased low level chronic inflammation (inflammaging)^10^, a decline in immune function^11^, and immunosenescence^12^. Inflammaging, specifically elevated plasma levels of innate inflammatory cytokines (including IL6 and TNF-α) have been independently associated with increased morbidity, frailty, and mortality in aging adults^13-15^.

Understanding the contributions of immunosenescence and inflammation to frailty will help to design preventative measures and therapies for frailty and promote healthy aging. Here, we investigate the potential link between soluble inflammatory indices, T cell expression of exhaustion and senescence markers, and frailty as measured by the VACS 2.0 index.

## MATERIALS AND METHODS

### Study Subjects

Informed consent was obtained from all subjects involved in the study, and studies were approved by the Cleveland VA Medical Center Institutional Review Board (IRB) or the Cleveland University Hospitals IRB.

Participants were excluded if they were HCV or HIV positive, had any acute viral illness, had immunological disease or immune modulatory treatments within 24 weeks of blood draw, or were pregnant or breastfeeding. Enrollment and a one-time blood draw occurred between the years 2017-2019, and health record data collection extended through May 5^th^, 2025. All participants enrolled were age 65 years and older, were male Veterans recruited from The Northeast Ohio VA Medical Center in Cleveland, Ohio. A small control cohort of younger participants was also enrolled for comparisons shown in figure 1 only. The majority (85%) of participants age 50 or younger, were non-veterans recruited from Case Western Reserve University in Cleveland, OH and were consented under either the Cleveland VA Medical Center IRB or the University Hospital IRB protocol. Depending on sample availability in Figure 1, n= 27-39.

**Figure 1.**
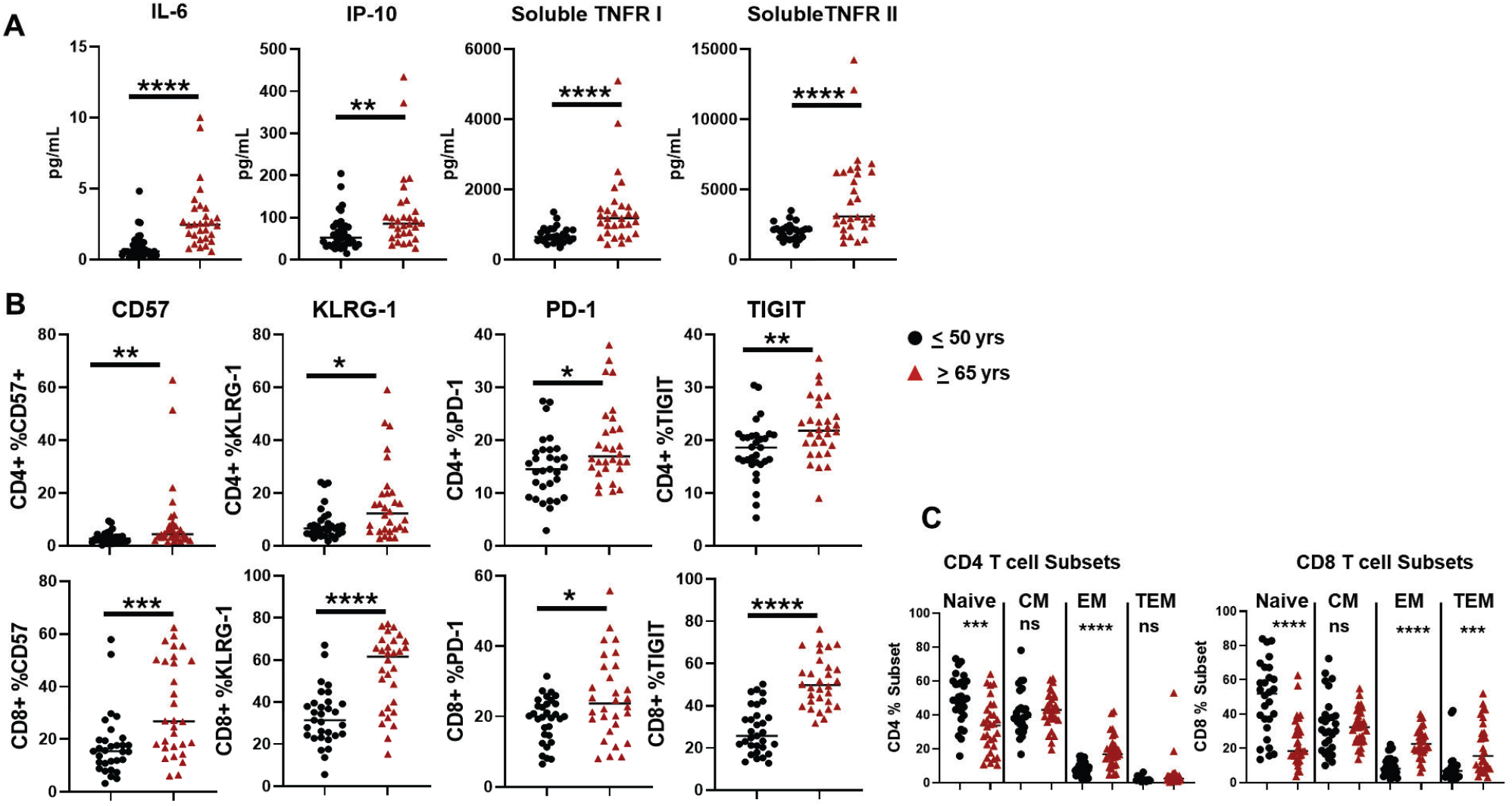
Plasma immune indexes and T cell markers of exhaustion and senescence are elevated in older Veterans. Plasma levels of inflammatory indices and T cell subsets (maturation, exhaustion, senescence) were compared between our older Veteran cohort (n= 30; 65 years and older) and a group of younger participants, (n=27-39; 50 years or younger). A) Plasma levels of soluble immune analytes (IL-6, IP10, sTNFRI, sTNFRII) were measured by ELISA. B) PBMCs were stained for T cell subsets and markers of senescence (CD57, KLRG-1) or exhaustion (PD-1, TIGIT). Live gated CD3+CD4+ or CD3+CD8+ T cells were assessed for proportion of CD57, KLRG-1, PD-1 or TIGIT expressing cells. C) Proportion of naïve (CD45RA+CD27+), central memory (CM) (CD45RA-CD27+), effector memory (EM) (CD45RA-CD27-), or terminal effector memory (TEM) (CD45RA+CD27-) CD4+ or CD8+ T cells were quantified. Red triangles ≥65 years old; black circles ≤50 years old; *p = <0.05; **p = <0.01; ***p = <0.001; ****p= <0.0001

All Veteran participant demographic, comorbidity, and pharmacological prescription data were obtained through the VA’s Electronic Health Record. Any active diagnoses or listed comorbidities within the year of the sample draw (6 months prior and 6 months post draw) were recorded. Any prescriptions listed as active medications 6 months prior, day of, or 6 months post sample draw were recorded. Prescribed medications alone were not used to assess comorbidity. Morbidities were only assessed if there was an active diagnosis in the health record. The number of prescriptions (Rx) were determined by the total number of active medications each participant was taking for the year surrounding the sample draw.

### Sample processing and storage

Participants donated a one-time blood sample. Plasma and serum were aliquoted and stored at -80°C. PBMCs were prepared from heparinized whole blood by ficoll-paque density sedimentation, frozen at -80°C and stored in liquid nitrogen until use. Samples were thawed and tested in batches containing roughly equal numbers of participants from each group (young and old, Figure 1).

### VACS 2.0 Frailty Score Calculation

We assessed frailty using the VACS 2.0 index, a composite score that combines demographic, HIV-related, and organ-system injury indicators. In our cohort of older, HIV uninfected patients, we modified the entry variables slightly as described here. The VACS 2.0 score includes the following variables: chronological age, sex, body mass index (BMI), CD4+ cell count, HIV-1 RNA viral load, hepatitis C virus (HCV) co-infection status, estimated glomerular filtration rate (eGFR), hemoglobin, alanine transaminase (ALT), aspartate aminotransferase (AST), platelet count, white blood cell (WBC) count, and albumin level (g/dL). In this cohort of older Veterans, those with current or previous HCV or HIV infection were excluded from enrollment. Therefore, zero was entered for HIV-1 RNA viral load and “no” was entered for the question of HCV co-infection status. We used the online VACS 2.0 tool, http://www.mdcalc.com/calc/10402/veterans-aging-cohort-study-vacs-2.0-index, to calculate VACS 2.0 frailty scores in our aging Veteran cohort.

### ELISA

Plasma IL-6 was measured by high-sensitivity ELISA (Quantikine HS, R&D Systems, Minneapolis, MN, USA), soluble tumor necrosis factor receptor (TNFR) I and II, and Interferon gamma-induced protein 10 (IP10) were measured by ELISA (Quantikine, R&D Systems, Minneapolis, MN, USA) following the manufacturer’s protocol. After the addition of the stop solution, plates were read on a VersaMax absorbance microplate reader and analyzed with SoftMax Pro version 5.4.1 software.

### Flow Cytometry

T cell phenotype was assessed using the following fluorochrome conjugated monoclonal antibodies: anti-CD3 PerCP (clone SK7), anti-CD57 FITC (clone NK-1) (BD Biosciences, San Jose, CA), anti-KLRG-1 APC (clone 13F12F2) (eBiosciences, San Diego, CA), anti-CD4 Pacific Blue (clone RPA-T4), anti-CD8 APCCy7 (clone SK1), anti-CD45RA PE-Cy7 (clone HI100), anti-CD27 AlexaFluor 700 (clone M-T271), anti-PD-1/CD274 BV711 (clone EH12.2H7), anti-TIGIT PE (clone A15153G) (Biolegend, San Diego, CA). PBMCs were incubated with viability dye (LIVE/DEAD-Aqua, Invitrogen, Grand Island, NY) at room temperature for 20 minutes then washed. Monoclonal antibodies were added for 20 minutes in the dark at room temperature, cells were then washed, fixed in PBS containing 2% formaldehyde, and events were acquired on a BD LSRFortessa flow cytometer (Becton Dickinson, San Jose, CA). Data were analyzed using FACSDIVA, (version 6.2 BD Bioscience, San Diego, CA) or FlowJo (version 10.5.0) software. T cell maturation subsets were determined based on expression of CD45RA and CD27; naïve = CD45RA+CD27+; central memory (CM) CD45RA-CD27+; effector memory (EM) CD45RA-CD27-; terminal effector memory (TEM) CD45RA+CD27-. If there were fewer than 100 events in any T cell memory subset, those data were excluded from analysis.

### Statistical Analysis

In Figure 1, comparisons between two unrelated groups (≤ 50 yrs vs ≥ 65 yrs) were performed using non-parametric two-tailed Mann Whitney U tests using GraphPad version 6 with significance thresholds set at p-values <0.05.

Plasma immune indices and T cell subset measurements were analyzed from frozen plasma or PBMCs. The clinical variables gathered from the Veteran patient records were collected as near to the time of blood draw as possible (on average, within 6 months of blood draw). A retrospective longitudinal follow-up analysis of patient charts was conducted to assess mortality and COVID-19 vaccine status.

We used a linear regression of chronological age (x-axis) versus VACS 2.0 score (y-axis) to categorize the older Veteran cohort as expected VACS 2.0 frailty if the value was within the 95% confidence interval, lower-than-expected VACS 2.0 frailty if the value fell below the 95% confidence level, or higher-than-expected VACS 2.0 frailty if the value fell above the 95% confidence level (Figure 3).

Correlations of VACS 2.0 score, age, absolute lymphocyte count (ALC), red blood cell distribution width (RDW), soluble immune indices, T cell subsets, and T cell markers of exhaustion and senescence were assessed using Spearman’s correlation and presented in a correlation heatmap, indicating correlations with Benjamini-Hochberg adjusted p-values < 0.05 (Figure 2).

**Figure 2.**
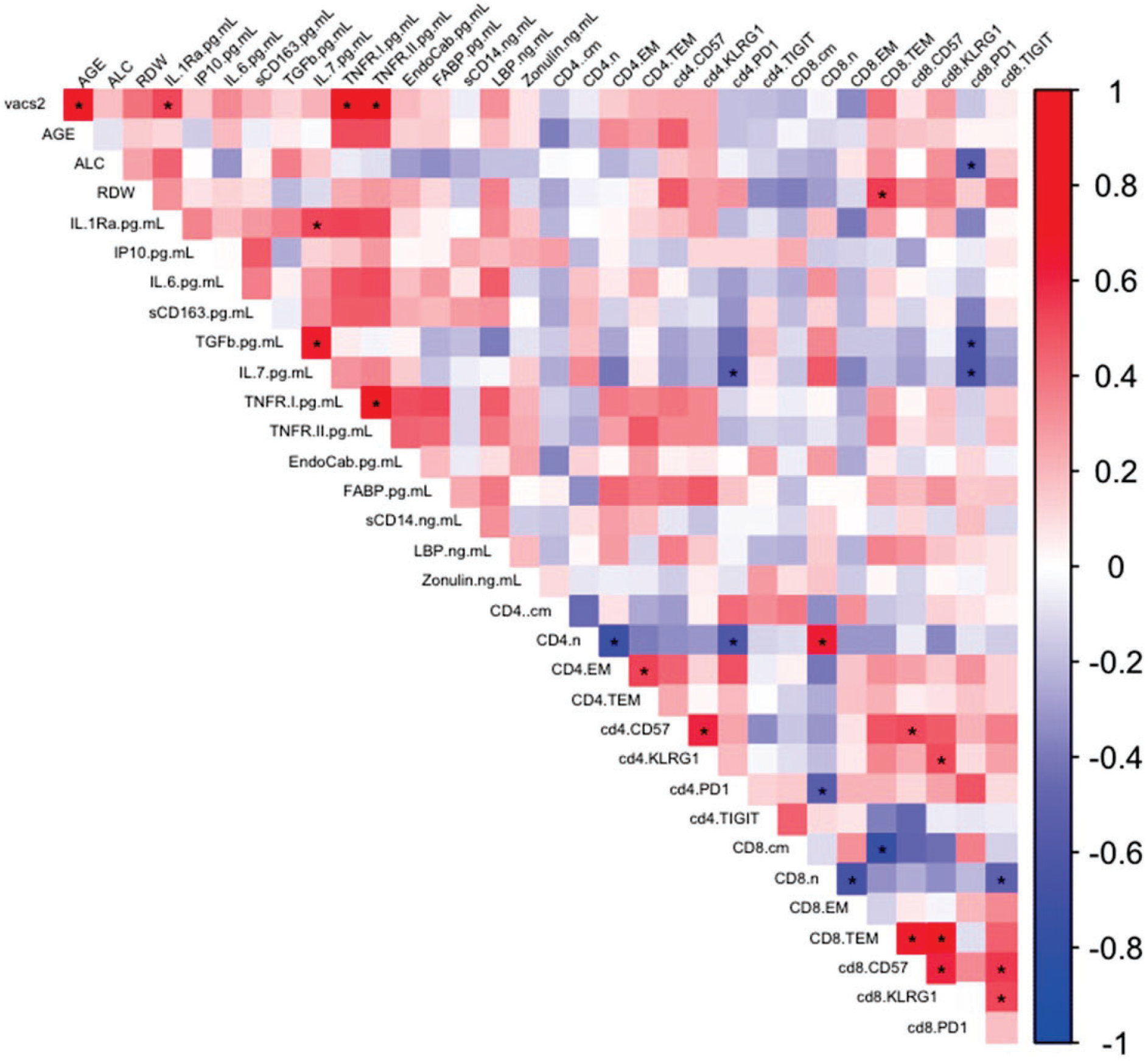
Spearman’s correlations of plasma immune indexes, T cell markers of exhaustion and senescence, age, absolute lymphocyte counts, and VACS 2.0 frailty score. Heat map showing Spearman’s correlation of VACS 2.0 frailty score, age, ALC, RDW, soluble immune indices, proportions of CD4+ or CD8+ naïve, CM, EM, or TEM T cells, and CD4 or CD8 T cell proportion of CD57, KLRG-1, PD-1, or TIGIT positive cells after adjustment for multiple comparisons. Red indicates positive correlation, blue indicates negative correlation, and color intensity represents “r” value of correlation. *significant correlation p= ≤0.05 Absolute lymphocyte count (ALC); red blood cell distribution width (RDW) For a full list of abbreviations on the heat map see Supplemental Table I.

## RESULTS

### Patient Characteristics

Our aging cohort of male Veterans 65 years or older ranged in age from 67 to 89 years and the average age was 77 years. They were 80% white and 20% black. Thirteen percent were current smokers, and the most common comorbidities were hypertension (87%), hyperlipidemia (57%), diabetes (30%), and coronary artery disease (27%) (Table 1). Thirteen percent of participants were taking 0 or 1 prescription, 30% were taking 2-4 prescriptions, 40% were taking 5-9 prescriptions, and 17% were taking 10 or more prescriptions (Table 1). The VACS 2.0 frailty scores ranged from 53-92, with a mean score of 66.

**Table 1.** Veteran Patient Characteristics.

| Category | Sub-Category | Frequency (N/total)<br>Percentage (%) |
| --- | --- | --- |
| Sex | Male | 30/30 (100%) |
|  | Female | 0/30 (0%) |
| Age | 65-79 | 19/30 (63%) |
|  | 80-89 | 11/30 (37%) |
| Race and Ethnicity | White non-hispanic | 24/30 (80%) |
|  | Black non-hispanic | 6/30 (20%) |
| Currently Smoking | Yes | 4/30 (13%) |
|  | No | 26/30 (87%) |
| Comorbidities | Diabetes | 9/30 (30%) |
|  | Hypertension | 26/30 (87%) |
|  | Hyperlipidemia | 17/30 (57%) |
|  | Coronary Artery Disease | 8/30 (27%) |
|  | Congestive heart failure | 4/30 (13%) |
|  | Myocardial infraction | 1/30 (3%) |
|  | Cardiomyopathy | 1/30 (3%) |
|  | Stroke, Hemorrhagic,<br>Ischemic Infraction or<br>unknown | 5/30 (17%) |
|  | COPD | 3/30 (10%) |
|  | Arthritis | 3/30 (10%) |
|  | Pulmonary Embolism/DVT | 2/30 (7%) |
|  | Chronic Kidney Disease | 5/30 (17%) |
| VACS 2.0 Index | 50-59 | 7/30 (23%) |
|  | 60-69 | 16/30 (54%) |
|  | 70-79 | 4/30 (13%) |
|  | 80+ | 3/30 (10%) |
| Polypharmacy | 0-1 Rx | 4/30 (13%) |
|  | 2-4 Rx | 9/30 (30%) |
|  | 5-9 Rx | 12/30 (40%) |
|  | 10+ Rx | 5/30 (17%) |

As mentioned above, most of the younger participants (85%) were non-veterans and we did not have access to their health records. The average age of the young participants was 30 years and ranged from 23 to 43 years old. Seventy-eight percent were male and 92% were white.

### Plasma immune indexes and T cell markers of exhaustion and senescence are elevated in older Veterans

We compared soluble immune indices associated with frailty (IL-6, IP-10, sTNFRI, sTNFRII) in our older Veteran cohort to our younger (≤50 years) control cohort. We found significantly elevated levels of plasma IL-6 (p= <0.0001), IP-10 (p= 0.001), sTNFR I (p= <0.001), and sTNFR II (p= <0.0001) in our older Veteran cohort (Fig. 1A). We also measured the proportion of CD4+ and CD8+ T cells expressing senescent markers (CD57, KLRG-1) and exhaustion markers (PD-1, TIGIT). Our older Veteran cohort also showed higher proportions of CD57 (p= 0.002), KLRG-1 (p= 0.02), PD-1 (p= 0.02), and TIGIT (p= 0.005) on CD4+ T cells and CD57 (p= 0.001), KLRG-1 (p= <0.0001), PD-1 (p= 0.03), and TIGIT (p= <0.0001) on CD8+ T cells compared to those of the younger cohort (Fig. 1B). Lastly, we saw significantly lower proportions of naïve CD4+ and CD8+ T cells and significantly higher proportions of memory CD4+ and CD8+ T cells in our older Veteran cohort compared to the young control cohort (Fig. 1C).

### Soluble TNFRs strongly correlate with VACS 2.0 Frailty Index

We utilized the Spearman’s correlation matrix to compare plasma immune indexes, T cell subsets and markers of exhaustion and senescence, age, ALC, RDW, and the VACS 2.0 frailty index score. We found that in this cohort, sTNFR I and sTNFR II were both significantly associated with VACS 2.0 frailty score (sTNFRI adjusted p= 0.001, r= 0.743; sTNFRII adjusted p= <0.001, r= 0.781) (Fig. 2). VACS 2.0 frailty scores were also statistically correlated with age (p= <0.001, r= 0.69) and plasma levels of IL-1Ra (p= 0.037, r= 0.53) (Fig. 2). While not statistically significant, age was also positively correlated with sTNFRs (sTNFRI adjusted p= 0.107, r= 0.511; sTNFRII adjusted p= 0.107, r= 0.512). Also, age was positively associated with the proportion of CD4 T cells expressing CD57, and was negatively correlated with the percent of CD4 CM T cells (Fig. 2). Although plasma levels of IL-6 did not correlate with VACS 2.0 scores as expected, IL-6 levels were associated (though not significantly) with sTNFR I and II (r= 0.49, r= 0.51, respectively) (Fig. 2). We also found that VACS2.0 score correlated with the number of prescriptions (p= 0.017, r= 0.43).

Lastly, we observed associations among soluble indices and between soluble indices and several T cell subsets (Fig. 2). A number of expected correlations between T cells subsets were evident as well (e.g. CD8%TEM correlated with CD8% CD57, KLRG, TIGIT) (Fig. 2 and Supplemental Table I).

### Veterans with higher-than-expected VACS 2.0 frailty for their given age had lower survival probability

Frailty is a known, but reversable, symptom of aging. However, it is also recognized that factors other than aging contribute to frailty development. In Figure 3, we examine the linear regression of VACS 2.0 frailty scores vs chronological age. Those that fell within the 95% confidence level of this linear regression were categorized as having “expected frailty”, those falling above were categorized as having “greater-than-expected” frailty, and those falling below were categorized as having “lower-than-expected” frailty. Figure 3 shows that 14 of 30 participants fell within the expected VACS 2.0 frailty category, 9 of 30 participants had lower-than-expected VACS 2.0 frailty, and 7 of 30 participants had higher-than-expected VACS 2.0 frailty.

**Figure 3.**
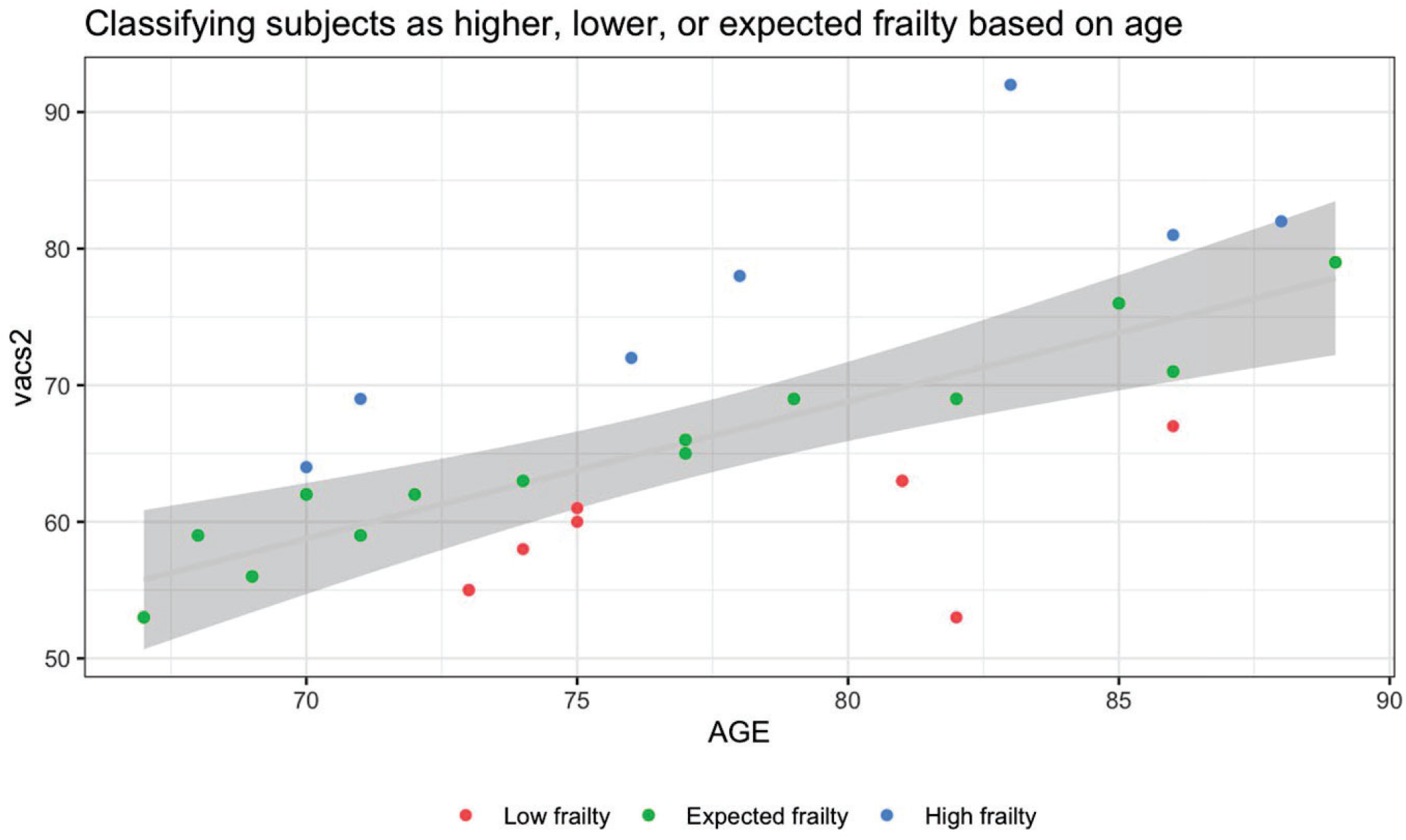
Identification of higher- or lower-than-expected VACS 2.0 frailty for given chronological age. The graph shows VACS 2.0 frailty score (y-axis) vs chronological age (x-axis) where each participant is represented by a scatter point. In the linear regression shown, any VACS 2.0 values falling within the 95% confidence level (grey) were categorized as “expected” frailty (n=14; green dots); those falling above the 95% interval were categorized as higher-than-expected frailty (n=7; blue dots); and those falling below the 95% interval were categorized as lower-than-expected frailty (n=9; red dots).

Interestingly, those patients categorized with “higher-than-expected” VACS 2.0 frailty had higher levels of sTNFRs and IP10, but not IL-6, when compared to patients with expected VACS 2.0 frailty (Supplemental Fig. 1). However, those in the “lower-than-expected” VACS 2.0 frailty category did not have lower levels of sTNFRs, IP10, or IL-6 when compared to patients with expected VACS 2.0 frailty. Not surprisingly, 4 of the 5 patients having 10+ prescriptions were categorized as having “higher-than-expected” VACS 2.0 frailty. Those with lower-than-expected VACS 2.0 frailty had a range in the number of prescriptions (0-8).

We further analyzed the predicted VACS 2.0 frailty categories and compared them to the observed mortality found in the patient health records since their enrollment in the study (Fig. 4). The time of study enrollment and sample draw was 2017-2019 and the last chart review was May 5, 2025. Of the 30 participants a total of 11 patients died, 5 were categorized as having higher-than-expected VAC 2.0 frailty, 3 were categorized as having expected VACS2.0 frailty, and 3 were categorized as having lower-than-expected VACS 2.0 frailty (Fig. 4). We observed that patients categorized with higher-than-expected VACS 2.0 frailty tended to have a lower survival probability compared to patients categorized in the expected or lower-than-expected VACS 2.0 frailty categories. Even in this small cohort, the differences in survival were nearly statistically significant (p= 0.057) (Fig. 4).

**Figure 4.**
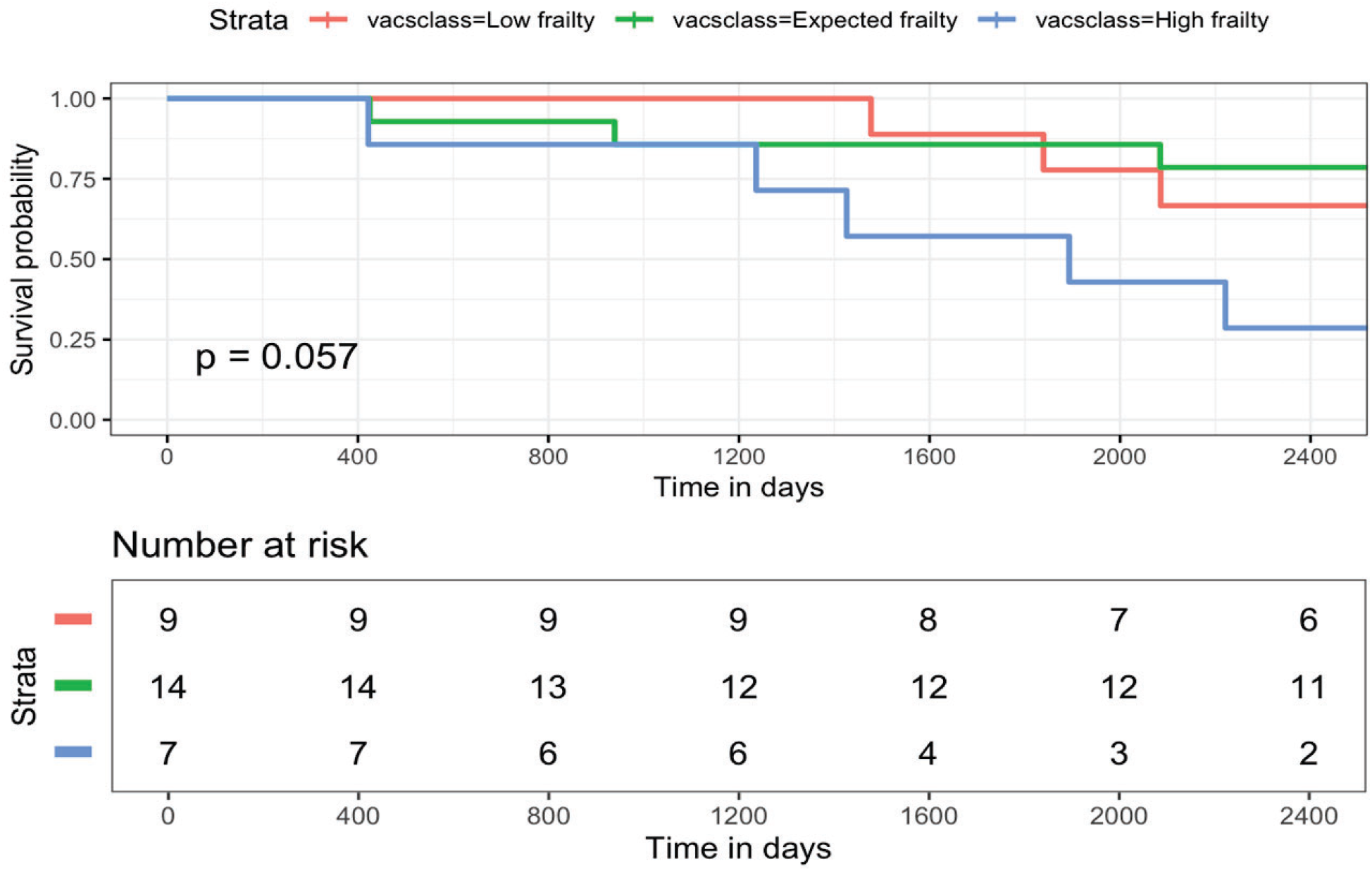
Patients with higher-than-expected VACS 2.0 frailty had a lower survival probability. The upper graph shows the survival probability of each of the VACS 2.0 frailty categories shown in figure 3 (expected, green line; higher-than-expected, blue line; lower-than-expected, red line). The time in days (x-axis) was calculated by counting the number days from patient sample blood draw to either recorded date of death or the last date of chart review (performed on May 5, 2025), whichever came first. The lower table shows the number of remaining participants in each of the 3 VACS 2.0 frailty categories over time (time in days).

## DISCUSSION

Frailty is, and will continue to be, a major health concern in the US and globally. Understanding the mechanisms behind the development of frailty will facilitate the design and the discovery of new therapies to promote a longer health-span.

The VA has the largest integrated health care and health records system in the US. Although not infallible, standardized procedures, clinical lab test, and standard of care are widespread (national) and fairly consistent. This allowed for the development of a frailty index that could be used across the US for Veterans. The Veteran Ageing Cohort was used to generate the first VACS index^16^ and was subsequently improved^5^. Although originally designed to examine frailty and predict 5-year mortality in older people with HIV, the VACS 2.0 index has since been validated in those without HIV^6, 7^. Of note, unlike other frailty indexes (e.g. The Veterans Affairs Frailty Index (VA-FI))^17^, the VACS 2.0 frailty index does not include diagnoses of neurocognitive deficiencies. However, the VACS index was found to associate with neurocognitive impairment^18^. In this study, contributions of morbidities related to neurocognitive defects, including depression, were not recorded. Also, because participants were excluded if taking immune modulatory treatments within 24 weeks of blood draw, it is likely that people with active cancer may have been excluded from the study.

Soluble inflammatory indices, specifically IL-6 and TNF, have been associated with mortality and frailty in older populations^13-15, 19^. Although we saw a highly significant elevation of plasma IL-6 and sTNFR I and II levels in Veterans ≥65 compared to the younger cohort (≤50yrs), only sTNFR I and II levels were correlated with higher VACS 2.0 frailty scores. One limitation of this study is the small cohort size, and is perhaps underpowered to see a significant correlation between plasma levels of IL-6 and the VACS 2.0 frailty scores.

A similar study was performed at the same VA Medical Center in Cleveland Ohio approximately 10 years earlier^20^. They analyzed frailty using the Fried frailty phenotype in a cohort of 117 Veterans ranging in age 62-95 years with a median age of 81 years. Using the Fried frailty phenotype, they were able to categorize the participants into non-frail, pre-frail, and frail. They found significantly higher levels of plasma IL-6, sTNFRI, and sTNFRII in the frail Veterans compared to the non-frail Veterans^20^. Also, they found that inflammation was more strongly associated with frailty than age. In our study, we also found significant correlations between VACS 2.0 score and plasma levels of sTNFRI and II (adjusted p= 0.001, r= 0.74; p= <0.001, r= 0.78, respectively) that was stronger than the correlation of age and plasma levels of sTNFRI and II (sTNFRI adjusted p= 0.107, r= 0.511; sTNFRII adjusted p= 0.107, r= 0.512). The previous study did not look at how Fried frailty related to T cell markers of exhaustion or senescence. In our current study, we found that those with higher-than-expected VACS 2.0 frailty had higher levels of plasma sTNFRs and IP10 and were more likely to have a greater number of prescribed medicines. Interestingly, the participants with lower-than-expected VACS 2.0 frailty did not show differences in soluble analytes, cell proportions or T cell expression of exhaustion/senescence markers compared to those with expected VAC 2.0 frailty. Nor did they show a greater or lower number of prescribed medications. One limitation of this study is the small cohort size and limited diversity. In Figure 1, 85% of participants in the young category were non-veterans for whom we had very little clinical data. The older Veteran cohort (n=30), while representative of the Cleveland VA Geriatric Clinic population, was all male. In general, females are more likely to be frail and the results from this study are not representative of the general public. A larger sample size may reveal other soluble marker or cell subset correlations that this small older Veteran cohort did not reveal.

Despite the limited cohort size, we found that those patients with higher-than-expected VACS 2.0 frailty tended to have a lower survival probability over time (Fig. 4). The post-sampling chart review extended 7-8 years (until May 2025), and overlapped with the 2019 COVID pandemic. We were able to collect participant chart data regarding COVID-19 vaccination and mortality. In this cohort of Veterans, 20/30 had a COVID-19 vaccine at some time between May 2020 and May 2025. COVID-19 infection may have exacerbated some of the participant’s existing comorbidity severity and it is likely that frailty impacted mortality from COVID-19 infection, however other risk factors cannot be rule out.

Here, we investigate the potential link between soluble inflammatory indices, T cell expression of exhaustion and senescence markers, T cell differentiation subsets, and frailty as measured by the VACS 2.0 index.

Overall, the results reveal that factors other than chronological age likely contribute more strongly to frailty as measured by the VACS 2.0 index. Inflammaging, particularly innate cytokines such as TNF, IL-6, and IL-1b, may contribute to frailty more than circulating T cell subsets. How and if these inflammatory indices, particularly sTNFRs, mechanistically contribute to frailty warrant future investigations.

**Supplemental Table 1:** List of abbreviations.

|  |  |
| --- | --- |
| VACS 2.0 | Veteran Aging Cohort Study |
| ALC | absolute lymphocyte count |
| RDW | red blood cell distribution width |
| IL-1Ra | interleukin-1 receptor antagonist |
| IP10 | Interferon-gamma-inducible protein 10 kDa |
| TGFb | transforming growth factor-beta |
| TNFR I | tumor necrosis factor receptor 1 |
| TNFR II | tumor necrosis factor receptor 2 |
| EndoCab | Endotoxin-core antibodies |
| LBP | lipopolysaccharide binding protein |
| FABP | fatty acid binding protein |
| cm | central memory |
| nav | naïve |
| EM | effector memory |
| TEM | terminal effector memory |
| KLRG1 | killer cell lectin-like receptor G-1 |
| PD1 | programed cell death protein-1 |
| TIGIT | T cell immunoreceptor with Ig and ITIM domains |

None of the authors or their institutions received any payments or services in the past 36 months from a third party that could be perceived to influence, or give the appearance of potentially influencing, the submitted work.

This research was supported by Veteran Affairs (VA) CDA grant 51K2CX001471 (Shive) and Merit grant BX006011 (Shive), and VA Merit BX1894 (Anthony); and by VA SHIELD funding support for Sarah Carbone.

## Data Availability

All data produced in the present study are available upon reasonable request to the authors

---

**Supplemental Figure 1.**
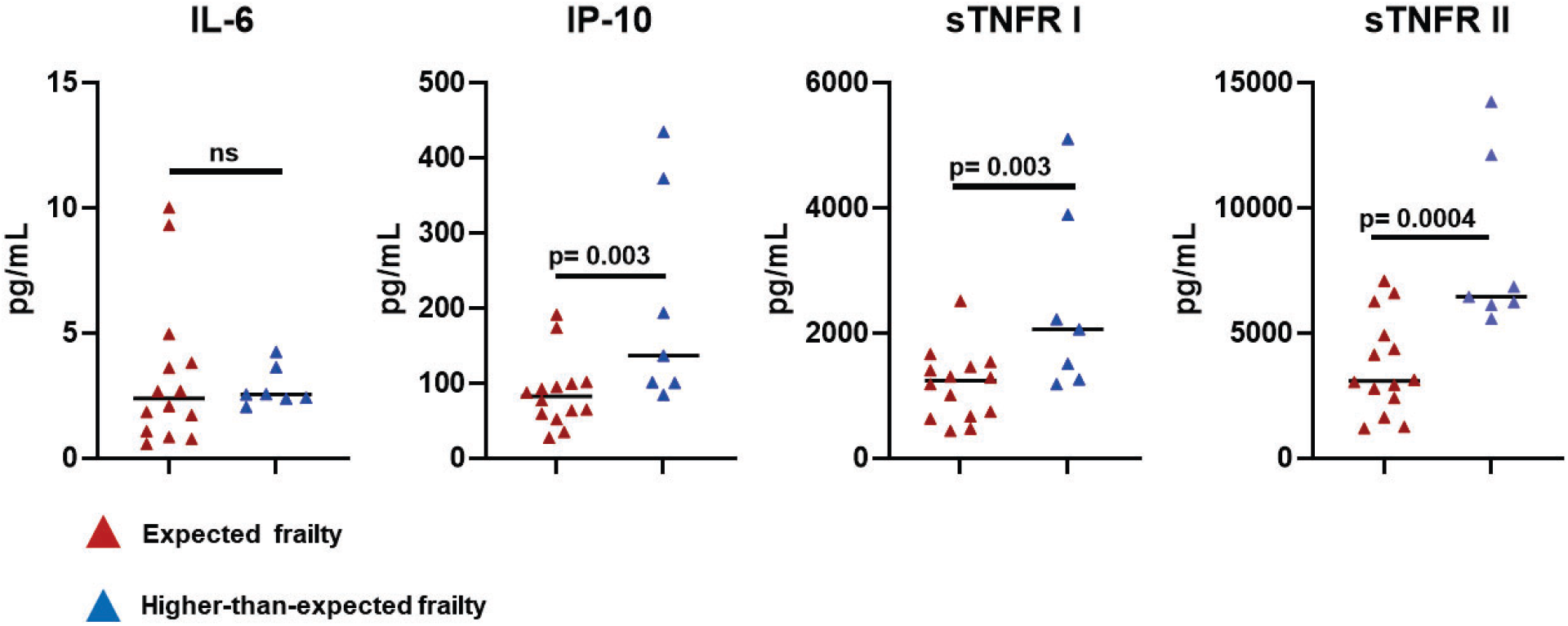
Soluble markers of inflammation are higher in older Veterans categorized as having “higher-than-expected” VACS 2.0 frailty scores. Plasma levels of soluble immune analytes (IL-6, IP10, sTNFRI, sTNFRII) from older Veterans categorized as having “higher-than-expected” VACS 2.0 frailty (n=7; blue triangles) were compared to those categorized as having “expected” VACS 2.0 frailty (n= 14; red triangles).

## CITATIONS

[1] Fried LP, Tangen CM, Walston J, Newman AB, Hirsch C, Gottdiener J, Seeman T, Tracy R, Kop WJ, Burke G, McBurnie MA, Cardiovascular Health Study Collaborative Research G: Frailty in older adults: evidence for a phenotype. The journals of gerontology Series A, Biological sciences and medical sciences 2001, 56:M146–56.

[2] Hoogendijk EO, Afilalo J, Ensrud KE, Kowal P, Onder G, Fried LP: Frailty: implications for clinical practice and public health. Lancet 2019, 394:1365–75.

[3] Bandeen-Roche K, Seplaki CL, Huang J, Buta B, Kalyani RR, Varadhan R, Xue QL, Walston JD, Kasper JD: Frailty in Older Adults: A Nationally Representative Profile in the United States. The journals of gerontology Series A, Biological sciences and medical sciences 2015, 70:1427–34.

[4] O’Caoimh R, Sezgin D, O’Donovan MR, Molloy DW, Clegg A, Rockwood K, Liew A: Prevalence of frailty in 62 countries across the world: a systematic review and meta-analysis of population-level studies. Age Ageing 2021, 50:96–104.

[5] McGinnis KA, Justice AC, Moore RD, Silverberg MJ, Althoff KN, Karris M, Lima VD, Crane HM, Horberg MA, Klein MB, Gange SJ, Gebo KA, Mayor A, Tate JP, North American ACCoR, Design a of the International Epidemiologic Databases to Evaluate A, Veterans Aging Cohort S: Discrimination and Calibration of the Veterans Aging Cohort Study Index 2.0 for Predicting Mortality Among People With Human Immunodeficiency Virus in North America. Clin Infect Dis 2022, 75:297–304.

[6] Akgun KM, Tate JP, Crothers K, Crystal S, Leaf DA, Womack J, Brown TT, Justice AC, Oursler KK: An adapted frailty-related phenotype and the VACS index as predictors of hospitalization and mortality in HIV-infected and uninfected individuals. J Acquir Immune Defic Syndr 2014, 67:397–404.

[7] Justice AC, Tate JP: Strengths and Limitations of the Veterans Aging Cohort Study Index as a Measure of Physiologic Frailty. Aids Res Hum Retrov 2019, 35:1023–33.

[8] Ganta N, Sikandar S, Ruiz SJ, Nasr LA, Mohammed YN, Aparicio-Ugarriza R, Cevallos V, Tang F, Ruiz JG: Incidence of Frailty in Community-Dwelling United States Older Veterans. J Am Med Dir Assoc 2021, 22:564–9.

[9] Justice AC, Dombrowski E, Conigliaro J, Fultz SL, Gibson D, Madenwald T, Goulet J, Simberkoff M, Butt AA, Rimland D, Rodriguez-Barradas MC, Gibert CL, Oursler KA, Brown S, Leaf DA, Goetz MB, Bryant K: Veterans Aging Cohort Study (VACS): Overview and description. Med Care 2006, 44:S13–24.

[10] Franceschi C, Bonafe M, Valensin S, Olivieri F, De Luca M, Ottaviani E, De Benedictis G: Inflamm-aging. An evolutionary perspective on immunosenescence. Ann N Y Acad Sci 2000, 908:244–54.

[11] Lopez-Otin C, Blasco MA, Partridge L, Serrano M, Kroemer G: The hallmarks of aging. Cell 2013, 153:1194–217.

[12] Pawelec G: Age and immunity: What is “immunosenescence”? Experimental gerontology 2018, 105:4–9.

[13] Harris TB, Ferrucci L, Tracy RP, Corti MC, Wacholder S, Ettinger WH, Jr., Heimovitz H, Cohen HJ, Wallace R: Associations of elevated interleukin-6 and C-reactive protein levels with mortality in the elderly. The American journal of medicine 1999, 106:506–12.

[14] Roubenoff R, Parise H, Payette HA, Abad LW, D’Agostino R, Jacques PF, Wilson PW, Dinarello CA, Harris TB: Cytokines, insulin-like growth factor 1, sarcopenia, and mortality in very old community-dwelling men and women: the Framingham Heart Study. The American journal of medicine 2003, 115:429–35.

[15] Michaud M, Balardy L, Moulis G, Gaudin C, Peyrot C, Vellas B, Cesari M, Nourhashemi F: Proinflammatory cytokines, aging, and age-related diseases. J Am Med Dir Assoc 2013, 14:877–82.

[16] Tate JP, Justice AC, Hughes MD, Bonnet F, Reiss P, Mocroft A, Nattermann J, Lampe FC, Bucher HC, Sterling TR, Crane HM, Kitahata MM, May M, Sterne JA: An internationally generalizable risk index for mortality after one year of antiretroviral therapy. AIDS 2013, 27:563–72.

[17] Orkaby AR, Nussbaum L, Ho YL, Gagnon D, Quach L, Ward R, Quaden R, Yaksic E, Harrington K, Paik JM, Kim DH, Wilson PW, Gaziano JM, Djousse L, Cho K, Driver JA: The Burden of Frailty Among U.S. Veterans and Its Association With Mortality, 2002-2012. The journals of gerontology Series A, Biological sciences and medical sciences 2019, 74:1257–64.

[18] Marquine MJ, Umlauf A, Rooney AS, Fazeli PL, Gouaux BD, Paul Woods S, Letendre SL, Ellis RJ, Grant I, Moore DJ, Group HIVNRP: The veterans aging cohort study index is associated with concurrent risk for neurocognitive impairment. J Acquir Immune Defic Syndr 2014, 65:190–7.

[19] Wikby A, Nilsson BO, Forsey R, Thompson J, Strindhall J, Lofgren S, Ernerudh J, Pawelec G, Ferguson F, Johansson B: The immune risk phenotype is associated with IL-6 in the terminal decline stage: findings from the Swedish NONA immune longitudinal study of very late life functioning. Mechanisms of ageing and development 2006, 127:695–704.

[20] Van Epps P, Oswald D, Higgins PA, Hornick TR, Aung H, Banks RE, Wilson BM, Burant C, Graventstein S, Canaday DH: Frailty has a stronger association with inflammation than age in older veterans. Immun Ageing 2016, 13:27.

